# Indirect Effects of COVID-19 on Maternal, Neonatal, Child, Sexual and Reproductive Health Services in Kampala, Uganda

**DOI:** 10.1101/2021.04.23.21255940

**Authors:** Jessica Burt, Joseph Ouma, Lawrence Lubyayi, Alexander Amone, Lorna Aol, Musa Sekikubo, Annettee Nakimuli, Eve Nakabembe, Robert Mboizi, Philippa Musoke, Mary Kyohere, Emily Namara, Asma Khalil, Kirsty Le Doare

**Affiliations:** School of Medicine, University of Leeds, Leeds, UK; Makerere University Johns Hopkins University, Kampala, Uganda; Maternal Vaccines Programme, Medical Research Council/Uganda Virus Research Institute at the London School of Hygiene and Tropical Medicine, Entebbe, Uganda; Department of Obstetrics and Gynaecology, Makerere University, Kampala, Uganda; Infection and Immunity, St. George’s, University of London, London, UK

**Keywords:** COVID19, maternal health, child health, vaccines, neonates

## Abstract

**Background:** COVID-19 impacted global maternal, neonatal and child health outcomes. We hypothesised that the early, strict lockdown which restricted individuals’ movements in Uganda limited access to services.

**Methods:** An observational study, using routinely collected data from Electronic Medical Records was carried out, in Kawempe district, Kampala. An interrupted time series analysis assessed the impact on maternal, neonatal, child, sexual and reproductive health services from July 2019 to December 2020. Descriptive statistics summarised the main outcomes before (July 2019 – March 2020), during (April 2020 – June 2020) and after the national lockdown (July 2020 – December 2020).

**Results:** Between 1^st^ July 2019 and 31^st^ December 2020 there were 14,401 antenatal clinic attendances, 33,499 deliveries, 111,658 childhood service attendances and 57,174 for sexual health. All antenatal and vaccination services ceased in lockdown for four weeks. During the three-month lockdown, the number of antenatal attendances significantly decreased, and remain below pre-COVID levels (370 fewer/month). Attendances for prevention of mother-to-child transmission of HIV dropped then stabilised. Increases during lockdown and immediately postlockdown included the number of women treated for high blood pressure, eclampsia and pre-eclampsia (218 more/month), adverse pregnancy outcomes (stillbirths, low-birth-weight and premature infant births), the rate of neonatal unit admissions, neonatal deaths and abortions. Maternal mortality remained stable. Immunisation clinic attendance declined whilst neonatal death rate rose (from 39-49/1000 livebirths). The number of children treated for pneumonia, diarrhoea and malaria decreased during lockdown.

**Conclusion:** The Ugandan response to COVID-19 negatively impacted maternal, child and neonatal health, with an increase seen in pregnancy complications, and fetal and infant outcomes, likely due to delayed care-seeking behaviour. Decreased vaccination clinic attendance leaves a cohort of infants unprotected, affecting all vaccine-preventable diseases. Future pandemic responses must consider impacts of movement restrictions and access to preventative services to protect maternal and child health.

**KEY QUESTIONS**

What is already known?

- The response to COVID-19 has been shown to have indirectly impacted on maternal, child, neonatal, sexual, and reproductive health.
- This is largely related to access to services and fear of contracting COVID-19 in outpatient departments.
- There has been very little data published on the health impacts of the COVID-19 response in Uganda.

What are the new findings?

- Antenatal attendances decreased dramatically in April, followed by increased numbers low-birthweight infants and neonatal deaths.
- Newborn immunisations against polio, tetanus, diphtheria, hepatitis B, haemophilus influenzae, rotavirus and pneumococcus decreased significantly.
- Sexual, and reproductive health services were reduced in number.

What do the new findings imply?

- Although Uganda has been less affected directly by COVID-19 infections in the first wave, the indirect impacts are far-reaching and will have future influences on population health.
- There is a degree of resilience within the healthcare service, but many services were adversely affected by the lockdown leading to poorer pregnancy and neonatal outcomes.
- Antenatal and vaccination services are of particular importance in ensuring the safety of mother and child and must be prioritised in the responses to future pandemics.

## INTRODUCTION

Uganda, as with many nations in the World Health Organization (WHO) Africa region, has largely avoided the considerable infection rate and death toll from COVID-19 that other nations saw during the first wave ^1 2^, with 38,085 confirmed cases and 304 deaths reported as of the 15^th^ January 2021 ^3^. Whilst this is likely under-representative of the true morbidity and mortality ^4^, Uganda has successfully minimised the spread and direct impact of COVID-19 within its borders through its early, rapid and severe response. However, maternal and child health services were severely impacted by these measures during the height of the lockdown, which may have indirectly affected morbidity and mortality in this group.

### Effects of COVID-19

Symptomatic COVID-19 infection in pregnancy is linked to worse maternal and neonatal outcomes than for pregnancies without COVID-19 ^5-7^. Studies across the UK and USA have also shown increased preterm birth, stillbirth, small for gestational age babies and neonatal mortality over the COVID-19 period and in relation to infection in pregnancy ^8^. However, global estimates of the indirect impacts of COVID-19 could amount to up to a 38.6% increase in maternal mortality, and 44.7% increase in child mortality per month across 118 low and middle income countries ^9^. The main factors proposed are disruptions to childbirth services and antenatal care (ANC) such as the management of pre-eclampsia and supplementation advice, wasting and curative child services,^9^ which the WHO have documented as being affected in many locations ^10^. Additionally, disruptions to family planning services including access to contraception and safe abortions, will result in an additional rise in maternal deaths, abortion-related complications, and a large unmet need for contraceptives ^11^. Further impacts on maternal and child outcomes may be seen through issues surrounding the provision of prevention and management of HIV ^9 12 13^, reduced lactation support ^14^ and conflicting guidance on whether to avoid breastfeeding if infected ^15^. These impacts have been reported in some low-resource settings ^10 16 17^, particularly with reduced antenatal attendances, linked to transportation restrictions, fear of transmission and lack of antenatal education ^17^.

### COVID-19 in Uganda

Preparation and readiness measures against COVID-19 in Uganda began between January and March 2020, focusing on health systems strengthening and capacity building, aided by early allocation of WHO funding ^18 19^. From the 2^nd^ March, the public were informed of the threat of COVID-19, with education and training subsequently disseminated ^18^. Testing focused on contacts of identified cases and those returning from travel, with population-wide lockdown measures imposed quickly after the first case in Uganda was reported on 21^st^ March 2020 ^20 21^. This included border closures, port-of-entry screenings, and quarantines for travellers ^18^. By the 25^th^ March this escalated to a ban on group-gatherings and non-essential internal travel, recommendation to work from home, and close schools ^18 22^. The travel restrictions included the cessation of all public transport and a ban on the use of private vehicles without explicit permission to travel ^23^. At a local level, non-essential visits to Kawempe National Referral Hospital (KNRH) were prohibited for a short time (23^rd^ March – 21^st^ April 2020), which included the closure of ANC and childhood immunisation clinics. The Ugandan Ministry of Health implemented PCR-based screening for symptomatic patients and any patient who was PCR positive was admitted to a dedicated ward. This paper aims to quantify the indirect impact of COVID-19 on maternal, neonatal and childhood outcomes at KNRH in Kampala.

## METHODS

This was a single-site observational study, which utilised retrospectively collected data, based in KNRH. This is a large, urban hospital with over 21,000 deliveries per annum, 200 antenatal clinic visits and 100 child admissions to hospital per day^24^. The hospital provides preventative and curative care during pregnancy and intrapartum, newborn and post-natal care, a paediatric ward and vaccination services at a standard indicative of care in urban Uganda. After the initial lockdown period (4 weeks without outpatient services), measures to reduce the number of women attending ANC included reducing the number of appointments per day from 150 to 90 for ANC and all women <26 weeks gestation being sent away to return after 30 weeks. For infants, the vaccination clinic remained operating routinely. During the initial phases of lockdown (April and May 2020) 35/60 doctors were reassigned to acute care at COVID centres in anticipation of a large number of COVID19 cases but 53 nurses were recruited at the same time with results-based financing support raising the number of nurse/midwifes on site from 184 to 237 after April 2020.

### Patient and Public Involvement

Patients were not involved directly in the formation of this study. We have involved women in a separate, dedicated qualitative study about their experiences of antenatal care during the pandemic^25^.

### Ethical Approval

This study received ethical approval from the School of Medicine Research Ethics (SOMREC 2020-148), Committee Uganda Council for Science and Technology (HS913ES).

### Data Collection

Data were retrospectively collected in January 2021, by hospital staff with access to the Electronic Medical Records (EMR) system. This system is part of the Uganda Ministry of Health (MoH) eHealth Policy, Strategy and Implementation Plan and utilises the District Health Information Software 2 (DHIS2) ^26^. The DHIS2 indicators for which data were collected are detailed in the Supplementary Materials and were taken from health management information system (HMIS) data, which is reported to the MoH, covering pregnancy preventative services, pregnancy curative services, childbirth, care of the newborn, postnatal care, preventative childcare, curative childcare, preventative services for women of reproductive age, curative services for women of reproductive age and unavailability of medicines and commodities. Monthly totals were gathered for the period of July 2019 to December 2020. In accordance with the Sex and Gender Equity in Research guidelines, pregnancy, childbirth and sexual health related indicators are reported for those of the female sex, and no segregation is made between male and female sex or gender for childcare indicators as this was not part of the reporting data ^27^.

Neonatal mortality was calculated as the sum of immediate neonatal deaths and deaths from neonatal Sepsis 0-7 days, neonatal sepsis 8-28 days, neonatal pneumonia, neonatal meningitis, neonatal jaundice, premature baby (as condition that requires management) and other neonatal conditions.

### Statistical Analysis

Data were input into Microsoft Excel and exported to R 4.0.4 (R Foundation for Statistical Computing, Vienna, Austria) for further analysis purposes. We calculated the number of attendances per month for each indicator then analysed the aggregated data at the month level as a proportion of the antenatal, labour and delivery, child health or sexual and reproductive health services attendance. For each indicator, the data were divided into pre-COVID (July 2019 – March 2020), lockdown (April – June 2020) and post-COVID lockdown (July – December 2020). We used descriptive statistics to summarise demographic and clinical data, and present summaries of outcomes before, during and after lockdown (the intervention) as medians and interquartile ranges (supplemental data 1). To identify suitable regression models for estimating the effects of lockdown, we first graphically plotted the number of events per month over time and assessed for stationarity using autocorrection using the Durbin-Watson test, graphs of residuals from ordinary least squares regression and graphs of auto and partial autocorrection functions. (supplemental figure 1). We then conducted interrupted time series analyses using the generalised least squares approach which allows for inclusion of autoregressive or moving average autocorrelation processes. These models were used to estimate the effects of lockdown on preventative, curative, labour and delivery and child health services at KNRH. The model included a time variable (month), a dummy lockdown variable indicating pre-, lockdown and post-lockdown, and trend variables for the lockdown and post-lockdown periods. This approach allows for estimation of the change in levels and trends of the outcomes following the multiple interruptions (start and lifting of lockdown). We use a 5% significance level and 95% confidence intervals (CI).

## RESULTS

Between 1^st^ July 2019 and 31^st^ December 2020 there were 14,401 ANC attendances, 33,499 deliveries, 111,658 attendances for childhood services and 57,174 SRH service attendances at KNRH. There was complete closure of all antenatal, sexual health and vaccination services for the first four weeks of the lockdown (23^rd^ March – 21^st^ April 2020).

### Pregnancy

#### Preventative Services

In the 9 months prior to lockdown at the end of March 2020, the median number of attendances for antenatal services was 894 per month (IQR 808-1035). During the three months of lockdown there were 539 fewer visits a month compared to pre-lockdown (95% CI 195.0-516.3, (p=0.001)). After the lifting of restrictions, the overall trend was for 370 fewer attendances compared to previous periods (95% CI 202.7-536.3, p=0.001).

The proportion of women receiving iron supplementation, tetanus vaccination and blood pressure monitoring remained unchanged after the initial closure of services at lockdown, despite fewer women attending ANC (Figure 1a-f). However, due to stockouts of intermittent antimalarial prophylaxis and folic acid supplementation prior to lockdown, there was an increase in the proportion of women receiving medication during lockdown.

**Figure 1.**
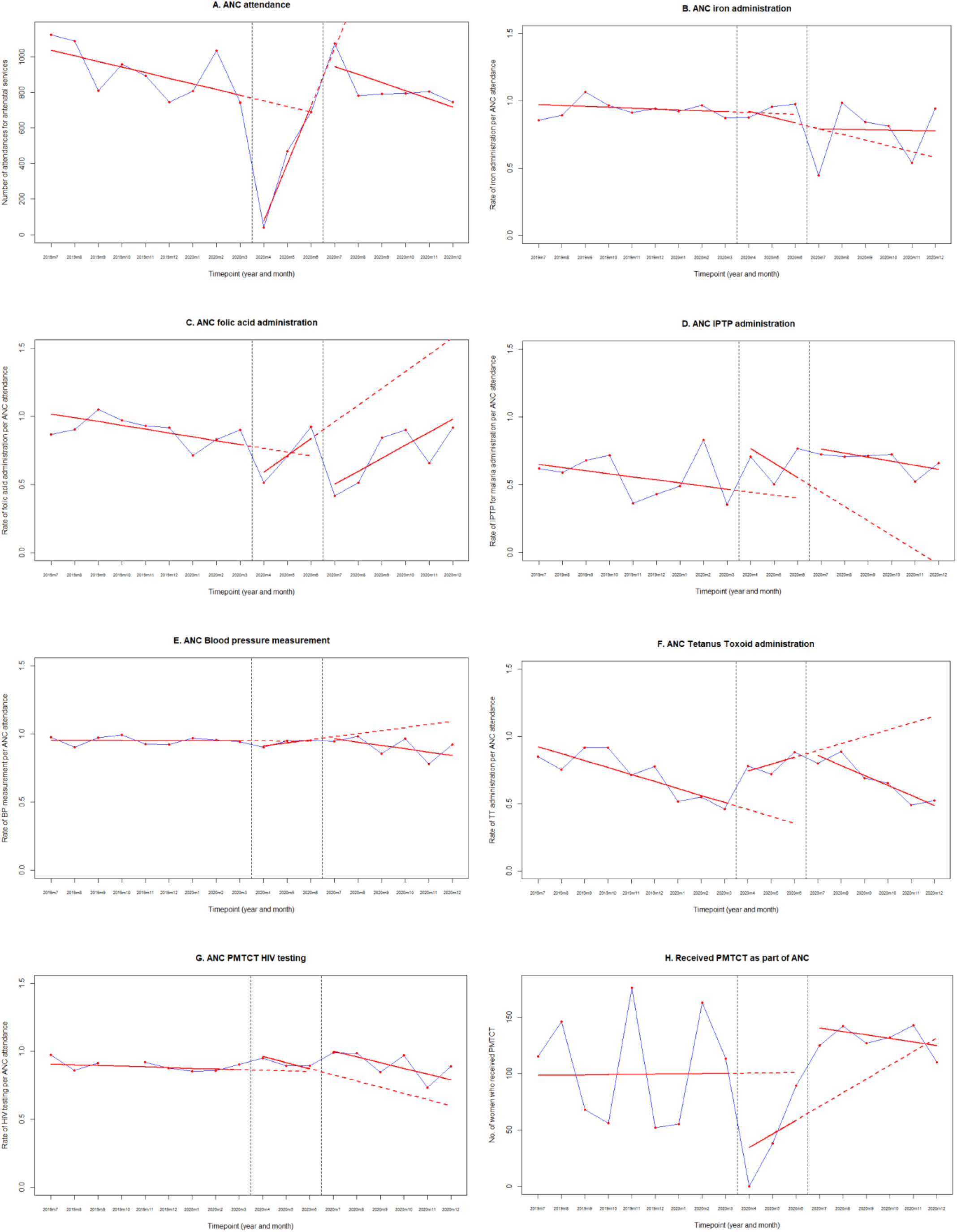
Interrupted time series analyses with generalised least squares regression for antenatal clinic services. A. antenatal clinic (ANC) attendance total attendances and rates of interventions as a proportion of total ANC attendance for: B. ANC iron administration, C. ANC folic acid administration, D. ANC intermittent antimalarial prophylaxis (IPTP) administration, E. ANC blood pressure measurement, F. ANC tetanus toxoid administration, G. ANC HIV testing. H. Number of women attending prevention of mother to child transmission of HIV (PMTCT) services as part of ANC, before, during and after the COVID-19 lockdown in Kawempe National Referral Hospital (KNRH). Red dots represent numbers/rates per month. Blue lines connect the red dots to display the observed monthly trend. Red solid lines represent the fitted regression models. Red dashed lines represent the counterfactual scenario.

The proportion of women receiving HIV testing in ANC declined by a rate of 4% (95% CI 1.5-6.5 decline p=0.001) during lockdown but increased back to baseline at the end of restrictions (figure 1g). The median number of women attending prevention of mother-to-child transmission of HIV services (PMTCT) before lockdown was 113 (IQR 56-146). Following the month of lockdown, during which time the clinic closed, the number of attendances increased slowly with a jump of 85 visits/month (95%CI 31.6-138.4, p=0.009) in the month that lockdown was lifted (Figure 1h).

#### Curative Services

The median number of women being treated for high blood pressure, pre-eclampsia and eclampsia prior to lockdown was 87/month (IQR 8-180). However, during the three months of lockdown there was a significantly increasing trend of 218 women/month (95%CI 108-327; p=0.002) receiving treatment. There has been a declining trend of 259 women/month (95% CI 153-365; p<0.001) receiving treatment in the months since lockdown was lifted. There was no change in the trend of women being treated for bacterial infections throughout the study, although this is incompletely captured in EMR.

### Labour and Delivery

The median number of monthly deliveries was 1869 (IQR 1791-1924) before March 2020 (pre-COVID). At the time of lockdown there were 320 more deliveries/month (95% CI 199-441, p=0.0002). During lockdown there was a trend of 109 (95% CI 55-163) fewer deliveries per month, although delivery trends have increased by a median of 117 (95% CI 54-180) deliveries per month since lockdown was lifted. During lockdown there was an increase in the rate of low birthweight infants (1.7% increase, 95% CI 0.6 – 2.7%, p=0.011) and in the immediate post-lockdown month an increase in stillbirths (1% increase, 95% CI −2% to 4%; p=0.58) and preterm births (6% increase, 95% CI −3%-15%; p=0.22), both not significant. There was no increase in the rates of maternal death over the course of the study (Figure 2a-g).

**Figure 2.**
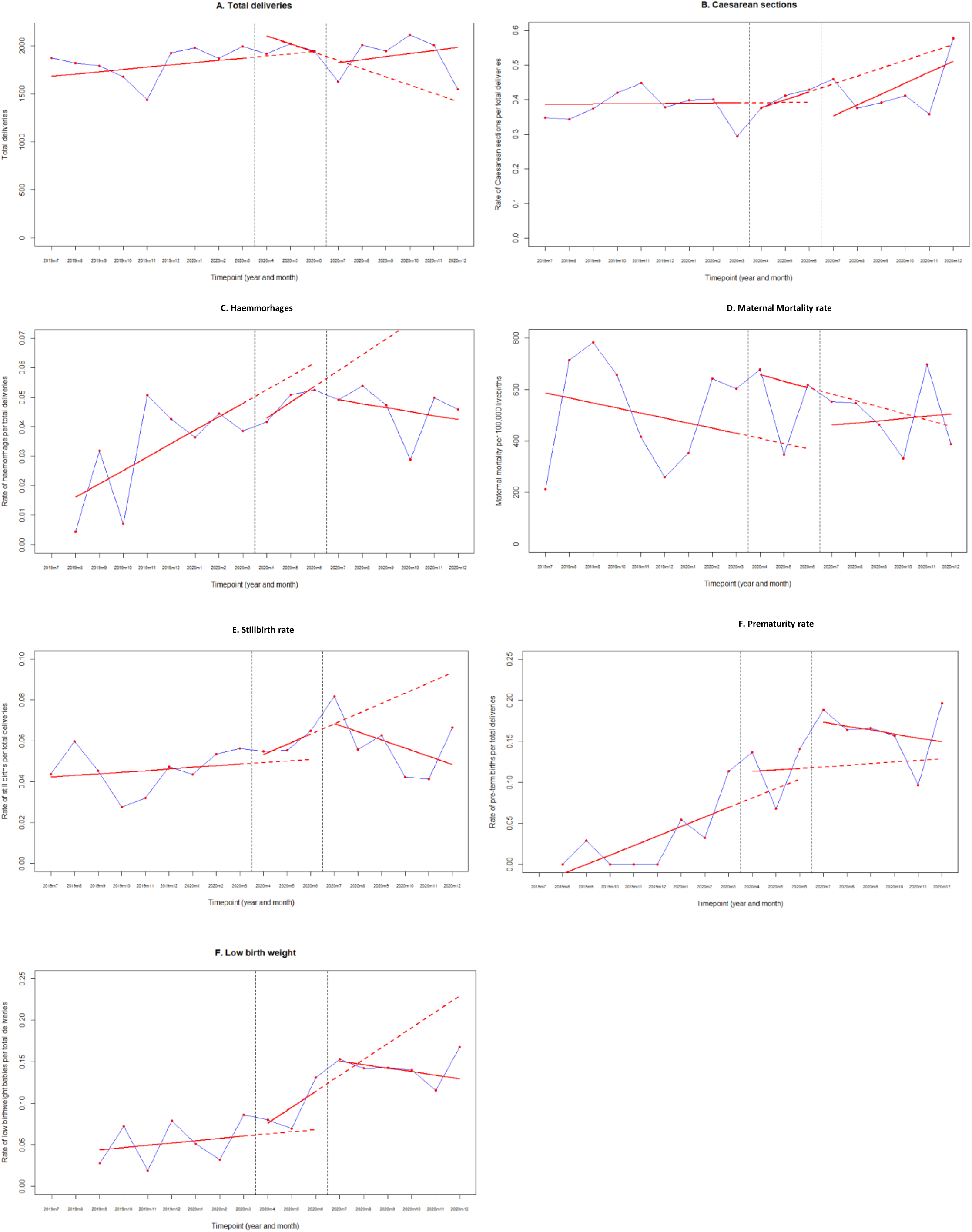
Interrupted time series analyses with generalised least squares regression for A. Total deliveries, B. Caesarean sections, C. Haemorrhage related to labour and delivery, D. Maternal mortality, expressed as rate per 100,000 maternities, E. Stillbirth rate as a percentage of deliveries, F. Preterm birth rate as a percentage of total livebirths, G. Low birth weight as a percentage of total livebirths, before, during and after the COVID-19 lockdown in Kawempe National Referral Hospital (KNRH). Red dots represent numbers/rates per month. Blue lines connect the red dots to display the observed monthly trend. Red solid lines represent the fitted regression models. Red dashed lines represent the counterfactual scenario.

### Care of the Newborn

Prior to lockdown, there was a median of 700 admissions per month to the neonatal unit (NICU) (IQR 652-706) and a neonatal mortality rate of 39.6/1000 livebirths (IQR 34.6-50.7). During lockdown there was an increasing rate of NICU admissions of 5.6% (0-11%; p=0.06). At the end of lockdown, the neonatal mortality rate increased by 10 neonatal deaths per 1000 livebirths/month (IQR 2-10; p<0.001) (figure 3a and 3b).

**Figure 3.**
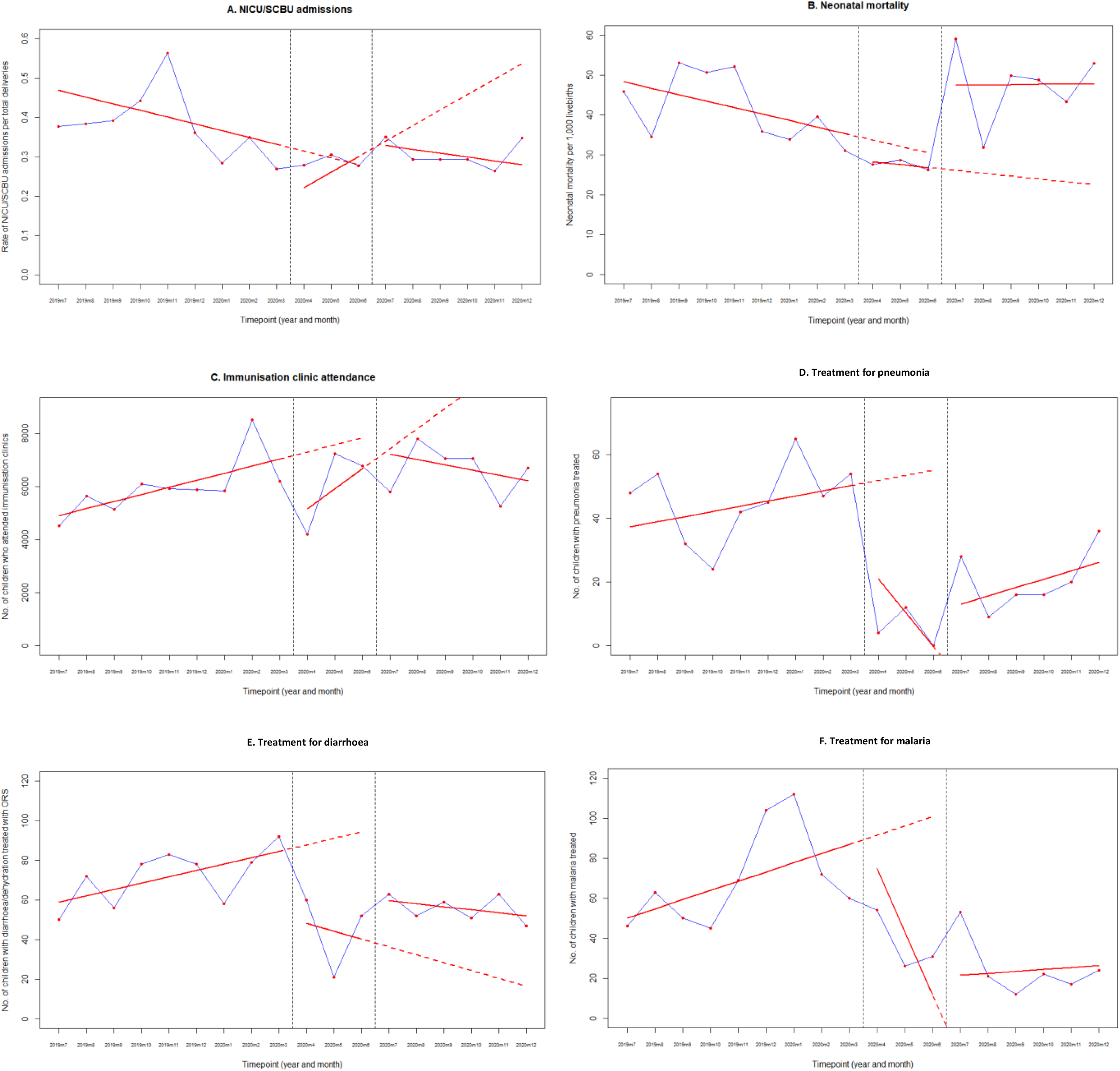
Interrupted time series analyses with generalised least squares regression for A. Neonatal intensive care unit admissions, B. Neonatal mortality rate expressed a rate per 1,000 livebirths, C. Immunisation clinic attendance, D. Inpatient Pneumonia treatment, E. Inpatient Diarrhoea treatment and F. Inpatient Malaria treatment, before, during and after the COVID-19 lockdown in Kawempe National Referral Hospital (KNRH). Red dots represent numbers/rates per month. Blue lines connect the red dots to display the observed monthly trend. Red solid lines represent the fitted regression models. Red dashed lines represent the counterfactual scenario.

### Post-natal care

The median number of women receiving immediate routine post-natal care services (within 24 to 48hrs of delivery) pre-COVID was 1873 (IQR 1823-1993). There were no cases of COVID-19 during this study.

### Child Health Services

Immunisations were offered on all 9 scheduled immunisation days in every month in 2019 and 2020. The median number of immunisation clinic attendances in the pre-COVID period was 5871 (95% CI 5643 – 6094). Since the lifting of lockdown there have been 960 fewer monthly attendances (771-2248; p=0.04)(Figure 3c). There was no change in the rate of children receiving BCG at birth, oral polio, pneumococcal or Rotateq vaccines since the end of lockdown, although fewer children now attend the immunisation clinic. The increase in the rate of measles vaccine is due to a catch up campaign after a long stockout (Supplementary figure 2).

There was a decline in the number of children being treated for pneumonia, malaria and diarrhoea (figure 3d-3f). There was an increase in the number of children treated for malnutrition after lockdown (supplementary table 1). We do not have data on infant or child mortality during the period.

### Sexual and Reproductive Health

There was a decrease in the number of women receiving the oral contraceptive pill (OCP) (13.6 more women; 95% CI 8.8-18.5 p<0.001) after restrictions were lifted. There were no changes to the number of women receiving intrauterine devices, although numbers recorded are small (Figure 4a and 4b). The number of sterilisation procedures and number of women treated for sexually transmitted diseases remained low both pre and post-lockdown. The number of abortions and sterilisations related to abortions increased by 338 (95% CI 58-619 more procedures; p=0.04) at the start of lockdown.

**Figure 4.**
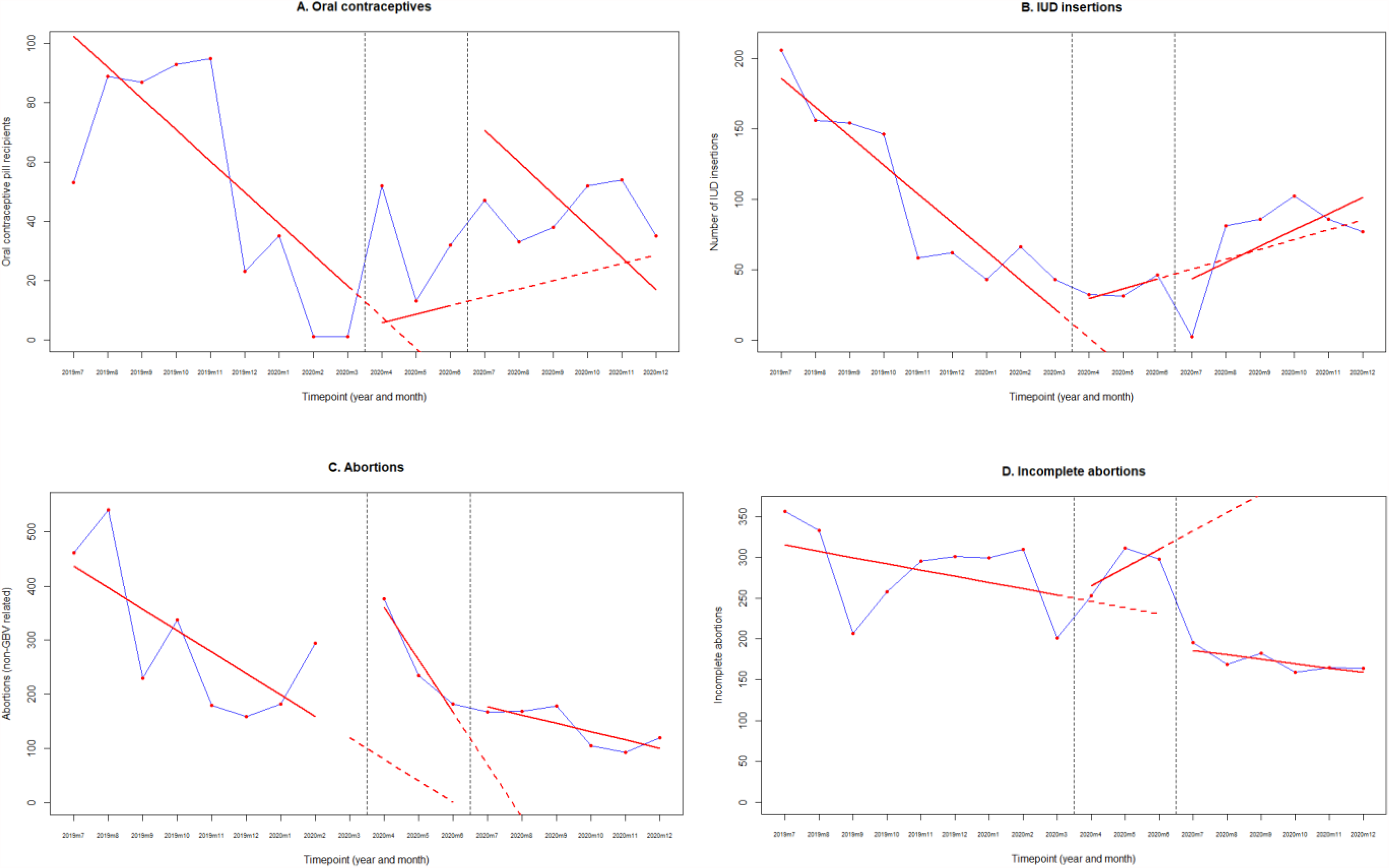
Interrupted time series analyses with generalised least squares regression for A. Oral contraceptive recipients, B. IUD insertions, C. Abortions and D. Incomplete abortions, before, during and after the COVID-19 lockdown in Kawempe National Referral Hospital (KNRH). Red dots represent numbers/rates per month. Blue lines connect the red dots to display the observed monthly trend. Red solid lines represent the fitted regression models. Red dashed lines represent the counterfactual scenario.

### Availability of Medicines

Several shortages were noted in medication and vaccination availability both pre- and post-lockdown which may have affected the ability to deliver effective services (Figure 5).

**Figure 5.**
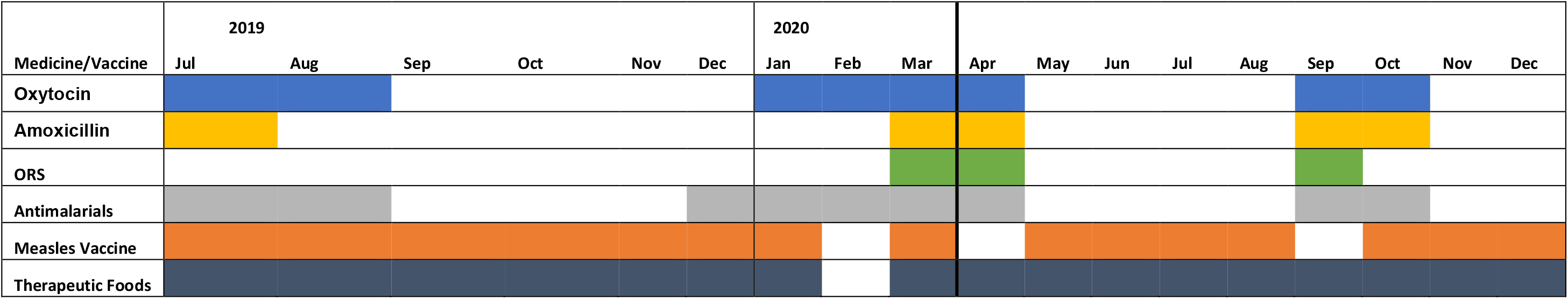
Overview of medicine stockouts by month. Coloured blocks indicate no stocks available, black lines indicate start and end of lockdown.

## DISCUSSION

Despite calls for the prioritisation of antenatal services and the consideration of the indirect impacts of lockdown restrictions on maternal health ^9 12 28^, the data from our study highlights that maternity, sexual and reproductive health, newborn and child health services were severely affected by COVID-19 restrictions.

Similar to our findings, facilities in rural Uganda saw a drop in antenatal attendances,^29^ as have hospitals in Kenya, Ethiopia, Zimbabwe and Rwanda in the first months of the pandemic ^30-37^. The aim of preventative services is to reduce maternal and newborn morbidity and mortality and any reduction in their availability can give an indication of the potential longer-term impacts – including increased rates of maternal anaemia, puerperal sepsis, stillbirth, low birth weight, preterm birth, malaria infection, pre-eclampsia/eclampsia, mother to child transmission of HIV and neonatal tetanus ^38^. Our study did not see an increase in maternal mortality despite fewer ANC attendances. This could be due to more women delivering in the community, however, our delivery rates remained constant throughout the period of study, suggesting that there may be alternative reasons for these findings, including increased maternal and neonatal morbidity, rather than mortality.

There are many proposed reasons why the ANC service and immunisation clinic attendances decreased so drastically during the lockdown. The national guidance at the start of the pandemic resulted in the closure of public transport, which a large proportion of patients rely on to access healthcare facilities, hence impacting their physical ability to access care, as has been reported in Uganda ^29 39^ and in other countries^30 34 36^. Other themes which have been reported to have affected attendances are the lack of healthcare staff, fear of infection, disruption of services due to COVID-19, lockdown orders restricting movement, and the increased price of transport ^33^. These themes have been highlighted in other studies in the region ^29 36 39^, indicating the need to consider the implications of lockdown measures on public confidence in healthcare in future emergencies. Whilst the number of ANC visits decreased, our delivery rate did not decline by the same amount. Kawempe hospital caters for a population of 2 million people, yet the number of women attending four ANC visits remains below 90%, although the majority of women in Kampala still deliver in hospital (94%)^40^. This data suggests that ANC and hospital delivery are not seen as a continuum of care in our setting and could account for the phenomenon of increased deliveries despite fewer ANC visits. Alternatively, fear of contracting COVID-19 in the community may have influenced the decision to give birth in a hospital environment.

The COVID-19 pandemic impacted on childbirth and deliveries across the region^5 41^, with some reports of decreased hospital deliveries ^30-32^. Facilities in Kenya have reported an increase in the number and rate of C-sections and fresh stillbirths ^34^, increased PPH ^31^ and an increase in the maternal deaths, disproportionately affecting adolescents ^34^. Whilst an increase in stillbirth and preterm birth was seen in our population at the end of lockdown, this was not significant. The age of the pregnant woman was not included in our dataset and may shed a valuable light on any population-level disparities in outcomes. Similar to other studies from Africa^30 32 36^, we advocate for prioritising safe and effective antenatal, intrapartum and postnatal care for future health emergencies.

The rise in neonatal deaths, low-birthweight babies and neonatal unit admissions are likely a result of the lack of antenatal care in March to May^38^. Sudden sharp changes in neonatal outcomes have been reported in South Africa, where an increase in neonatal mortality was linked to the disruption of services and diversion of resources due COVID-19 necessities ^42^. As seen with our data, a hospital in Malawi found an increase in babies born earlier and at lower birth weights, however the same study did not find this in a Zimbabwean hospital ^43^ suggesting there are differences between countries that remain unexplained.

A decrease in children attending hospital, as seen in our findings, was also seen in South Africa ^42^ and Ethiopia ^37^. This is likely also associated with fear of attending healthcare settings, inaccessibility, and a reduction in self-referrals, as seen with ANC ^37^. Conversely, there was an increase in malnutrition attendances, likely due to the societal impacts of COVID-19 restrictions on child health and nutrition ^44^. The lack of therapeutic foods available may have been affected by border closures and trade restrictions, in a similar manner to medication availability in Nigeria ^45^.

The reduction in immunisation clinic attendances in our cohort puts an estimated 20,000 children at risk of mortality from vaccine-preventable diseases such as tetanus and polio^46^. Uganda was declared polio-free in 2010 ^47^. However, the situation remains precarious due to the possibility of imported virus in surrounding countries where polio is not yet irradicated ^48^. The likelihood of a potential infectious disease outbreak can be predicted based on the proportion of coverage lost by a period of reduction in immunisations and The WHO estimates that at least 80 million children will be at risk of diseases like tetanus, polio, diphtheria and measles due to disruption of vaccination programmes during the pandemic ^49^. A reduction in the uptake of childhood immunisations were also seen in England ^50^ and Singapore ^51^ at the start of the pandemic, highlighting the universal impact of COVID-19 on child health. The follow-up response to vaccine catch-up in this pandemic is of key importance in mitigating future outbreaks and further impacts on child health ^52 53^.

Whilst our data shows some resiliency in sexual health and contraceptive services, the reduction in HIV clinic attendances is worrying and requires further outreach work to ensure the provision of care. PMTCT services bounced back quickly, in part due to international funding support mechanisms in place, such as that from USAID ^54^. Sexual and reproductive health services have also been impacted across other East African nations. Facilities in Kenya and Ethiopia reported contraceptive services were limited and decreases were seen in family planning attendances due to the closure of services ^32 37^, although national data from Kenya show no change overall in the usage of services ^34^.

### Clinical and Research Implications

Whilst the effects of COVID-19 are wide ranging, the continuation of changes to antenatal services, maternal and neonatal outcomes, and reduced number of children being treated for pneumonia and malaria in hospital through to December 2020 may also be influenced by other social factors. Many staff and patients were affected by restrictions to movement in October and November 2020 due to political campaigning and riots relating to the presidential elections. This highlights the susceptibility of health and healthcare services to wider events, reinforcing the need for resilience and planning going forward.

### Strengths and Limitations

Although the limitations of this study lie in the use of data from a single site, collected retrospectively, this has allowed the inclusion of over 33,000 births, 14,401 antenatal attendances and 111,658 childhood immunisations, highlighting the massive impact on this population. Furthermore, the use of data from EMR rather than direct patient records mean this data is likely an under-representation of the true values of each indicator. Statistical comparison using data from the full year of 2019 would have enabled a better understanding of how 2020 compared to the time before COVID-19. Even with these documented limitations, our findings reinforce the importance of considering maternal and child health in future pandemic responses.

## CONCLUSIONS

Maternal, neonatal, child, sexual and reproductive health services were all impacted by the restrictions imposed by the Ugandan government in response to COVID-19. Our results demonstrate the urgent need for pandemic responses to take into account the local context, where a stringent lockdown may be detrimental to the overall health of the population. Such responses must include the prioritisation of preventative care including maintaining antenatal clinic visits, child health and vaccination services to prevent delayed impacts on maternal, neonatal and child health. Furthermore, any disruptions to immunization schedules must be mitigated as rapidly as possible, to prevent further infectious disease outbreaks and future pandemics.

## Data Availability

Data will be available from the SGUL research repository, to be input

https://sgul.figshare.com/

## ACKNOWLEDGEMENTS

We would like to thank the electronic medical records team at KNRH for access to the data for this study.

