## Supplementary figures and images for "Indirect Effects of COVID-19 on Maternal, Neonatal, Child, Sexual and Reproductive Health Services in Kampala, Uganda"

### Supplementary Figure 1

Supplementary Figures 2 – Doses of childhood vaccines administered at KNRH, Jul19 to Dec20

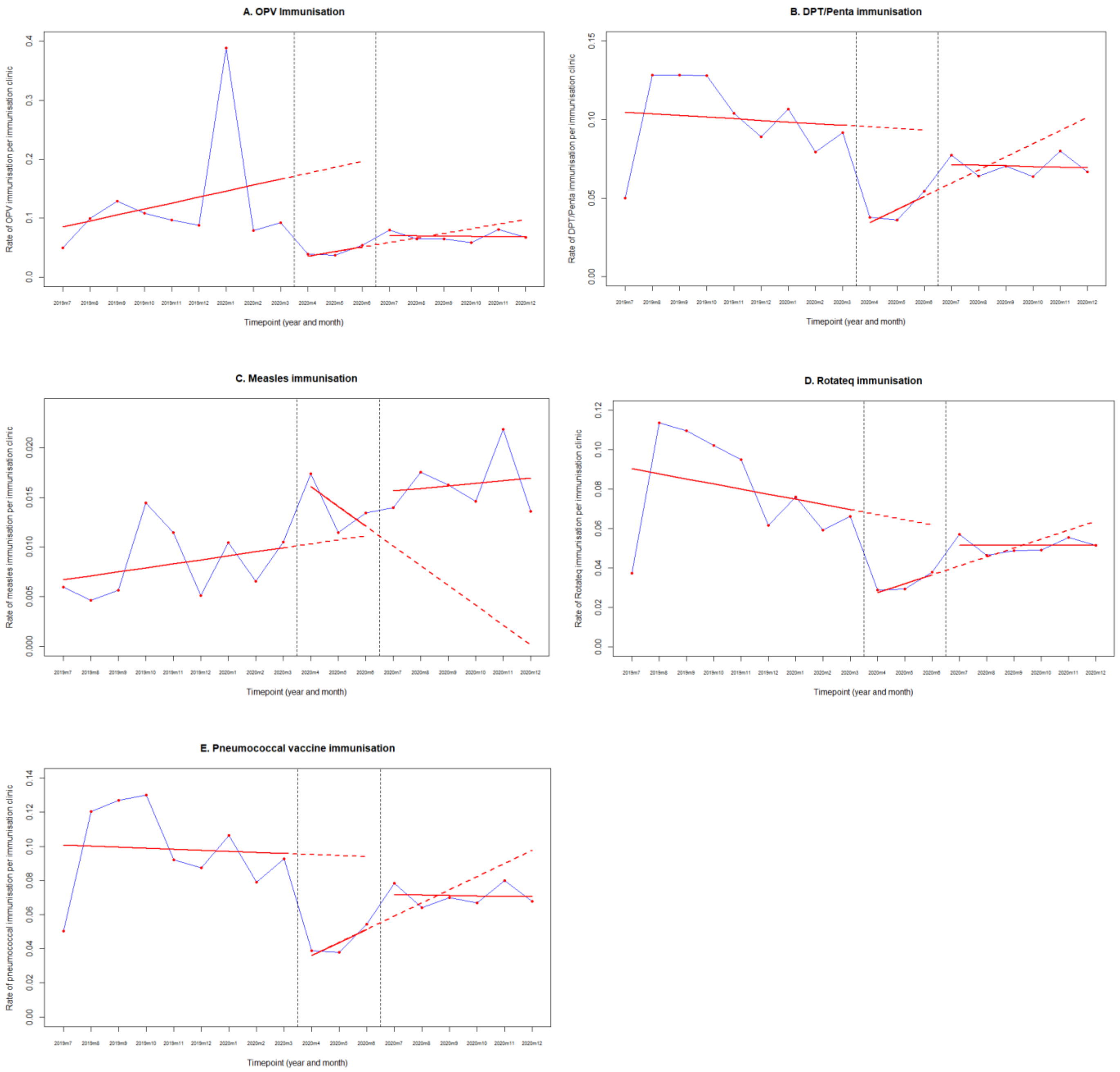

### Supplementary Figure 2

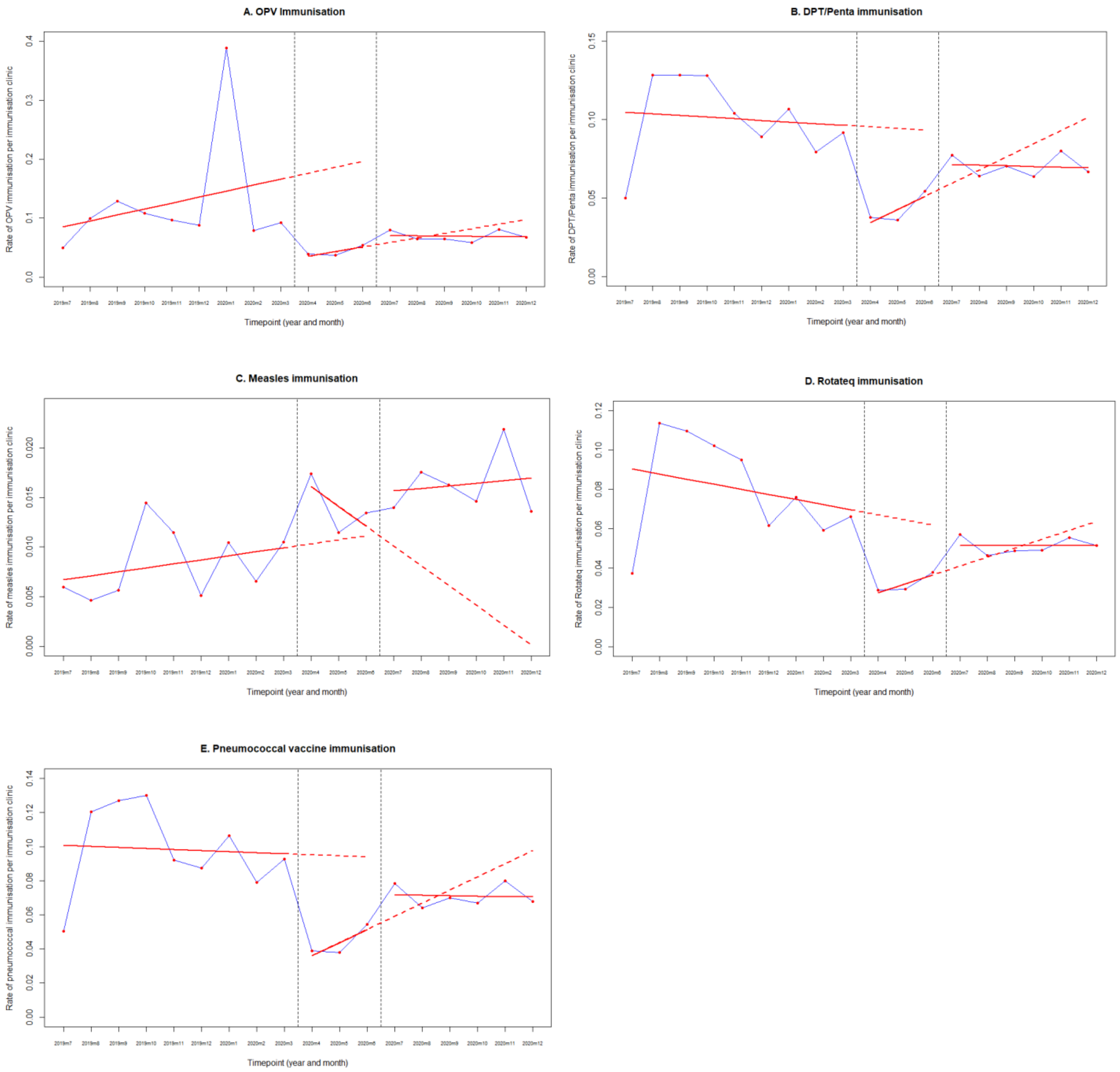
